# A comprehensive knowledgebase of known and predicted human genetic variants associated with COVID-19 susceptibility and severity

**DOI:** 10.1101/2022.11.03.22281867

**Authors:** Meltem Ece Kars, David Stein, Çiğdem Sevim Bayrak, Peter D Stenson, David N Cooper, Yuval Itan

## Abstract

Host genetic susceptibility is a key risk factor for severe illness associated with COVID-19. Despite numerous studies of COVID-19 host genetics, our knowledge of COVID-19-associated variants is still limited, and there is no resource comprising all the published variants and categorizing them based on their confidence level. Also, there are currently no computational tools available to predict novel COVID-19 severity variants. Therefore, we collated 820 host genetic variants reported to affect COVID-19 susceptibility by means of a systematic literature search and confidence evaluation, and obtained 196 high-confidence variants. We then developed the first machine learning classifier of severe COVID-19 variants to perform a genome-wide prediction of COVID-19 severity for 82,468,698 missense variants in the human genome. We further evaluated the classifier’s predictions using feature importance analyses to investigate the biological properties of COVID-19 susceptibility variants, which identified conservation scores as the most impactful predictive features. The results of enrichment analyses revealed that genes carrying high-confidence COVID-19 susceptibility variants shared pathways, networks, diseases and biological functions, with the immune system and infectious disease being the most significant categories. Additionally, we investigated the pleiotropic effects of COVID-19-associated variants using phenome-wide association studies (PheWAS) in ∼40,000 BioMe BioBank genotyped individuals, revealing pre-existing conditions that could serve to increase the risk of severe COVID-19 such as chronic liver disease and thromboembolism. Lastly, we generated a web-based interface for exploring, downloading and submitting genetic variants associated with COVID-19 susceptibility for use in both research and clinical settings (https://itanlab.shinyapps.io/COVID19webpage/). Taken together, our work provides the most comprehensive COVID-19 host genetics knowledgebase to date for the known and predicted genetic determinants of severe COVID-19, a resource that should further contribute to our understanding of the biology underlying COVID-19 susceptibility and facilitate the identification of individuals at high risk for severe COVID-19.

## Main

Investigating host genetic variation has been instrumental for understanding the pathophysiological processes that lead to severe COVID-19^1-4^. A substantial number of studies have explored the genetic factors underlying COVID-19 host susceptibility/severity using a variety of methods, including: (1) candidate gene approach and gene burden tests for identifying immune system disorders by means of whole-exome or whole-genome sequencing, or large-scale genome-wide association studies (GWAS); (2) frequency calculations of risk variants in population-based sequencing cohorts; (3) functional studies; and (4) various *in silico* methods such as molecular docking studies and binding affinity predictions ^2,5-9^. Consequently, the number of identified COVID-19-associated host genetic susceptibility variants has rapidly increased. However, current resources (along with review articles) include a relatively small set of variants^10-12^ and are mostly derived from GWAS summary results^4^.

COVID-19-associated variants have been identified by different methodologies, complicating the confidence assessment of these variants as pathogenic. Also, there is currently no comprehensive resource describing published COVID-19-associated variants, with no tools available to computationally predict novel COVID-19 severity variants other than general variant pathogenicity predictors. Generating an *in-silico* tool specifically tailored for predicting COVID-19 severity variants would facilitate the discovery of novel variants and contribute to the research on the pathogenesis of severe COVID-19.

To address the shortcomings in the research of COVID-19 host-genetics, we generated a comprehensive COVID-19 host genetics knowledgebase, presenting the first machine learning classifier of COVID-19 host genetic variants and precalculated predictions for all possible human missense variants based on a model that learned from a broad set of variant-, gene-, protein- and network-level features. We also explored the biological properties of the variants and genes implicated in COVID-19 susceptibility using feature importance, gene-set enrichment, pathway and network analyses and conducted PheWAS for identifying shared physiological processes of COVID-19 and diseases associated with severe COVID-19 (Fig. 1). Finally, we generated the COVID-19 Host Genetic Variants webpage, a comprehensive resource for the known and predicted COVID-19-associated variants.

**Fig. 1:**
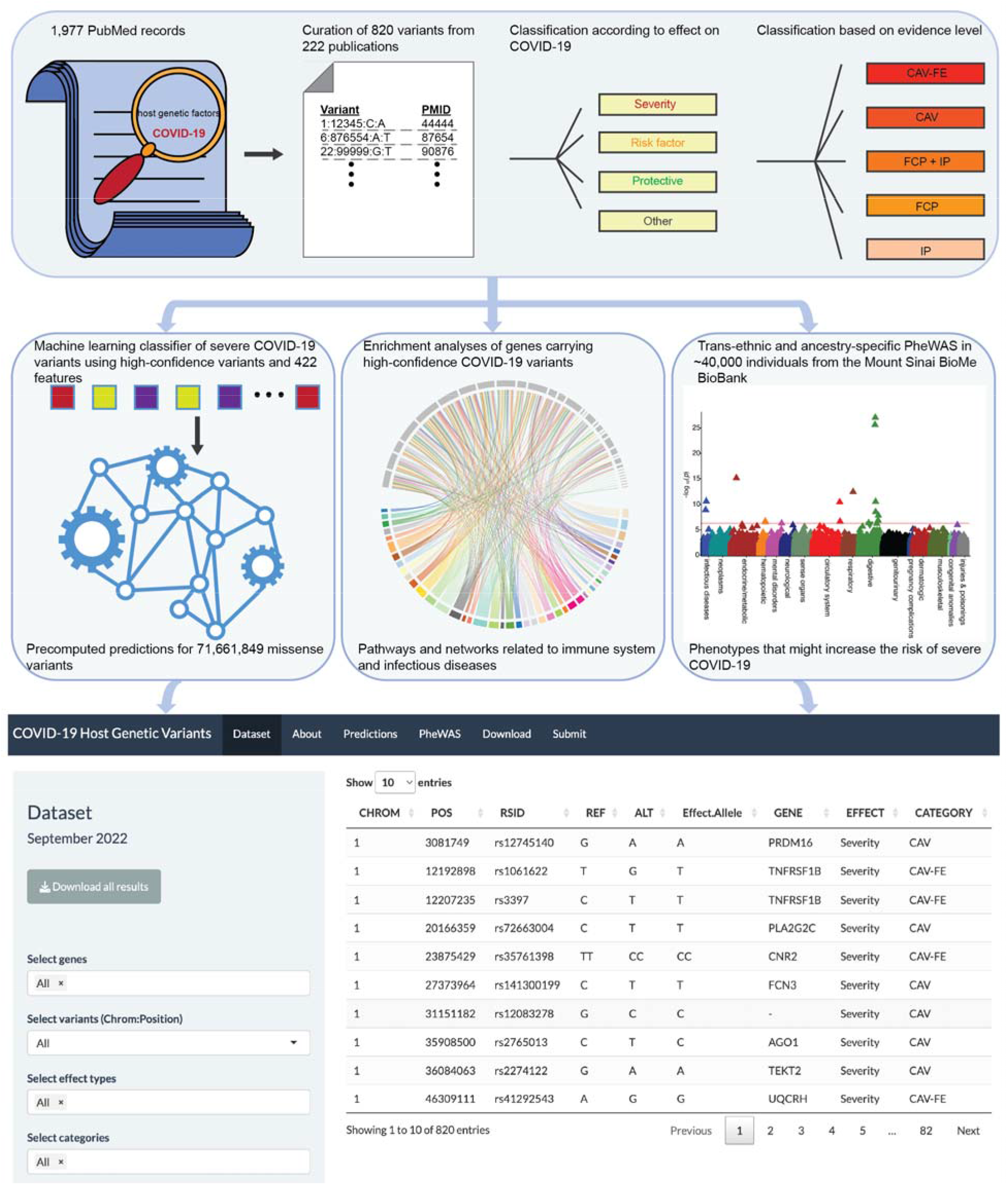
Overview of the study workflow. Panels depict the curation and categorization of COVID-19-associated host genetic variants (top), generation of a machine learning classifier of severe COVID-19 variants (middle left), pathway and gene-set enrichment analyses (middle center), PheWAS in the Mount Sinai BioMe BioBank (middle right), and the web-based interface of the COVID-19 Host Genetic Variants (bottom).

## Results

### Curation and categorization of COVID-19 host genetic variants

Through text mining 1,977 publications related to host genetic factors associated with COVID-19 susceptibility/severity and systematic manual curation of 222 eligible studies, we obtained a list of 820 COVID-19-associated host genetic variants of which 719 were located in 295 different genes whilst 101 were located within intergenic regions (Fig. 1, Methods). We categorized the COVID-19-associated variants into four groups: (1) protective, variants associated with asymptomatic or milder disease; (2) risk factor, variants suggested to increase the risk of symptomatic COVID-19; (3) variants known to be associated with a severe form of disease, e.g., admission to an intensive care unit and critical COVID-19 pneumonia; and (4) other effects, e.g., variants which destabilize the structure of proteins related to COVID-19 susceptibility even if their impact on the course of the disease is unknown. To obtain a list of high-confidence COVID-19-associated variants, variants were also categorized into five confidence levels based on the published evidence (Fig. 1 and Table 1). These categories include (1) COVID-19-associated variants with functional evidence (CAV-FE): variants identified through association studies or a candidate gene approach and where supporting functional data are available (e.g., gene expression, cell-based assay data), (2) COVID-19-associated variants (CAV): variants identified through association studies or a candidate gene approach, (3) Allele frequency - COVID-19 prevalence correlation (FCP): variants identified in studies that have investigated the relationship of the frequency of potential COVID-19-associated variants and COVID-19 prevalence in different populations. (4) *In silico* prediction (IP): variants that were identified as being deleterious in studies that used only *in silico* methods to predict the effect of amino acid exchange on COVID-19 susceptibility (e.g., docking, binding affinity predictions, structural modeling). (5) Allele frequency - COVID-19 prevalence correlation plus *in silico* prediction (FCP + IP): variants that fall into both FCP and IP categories.

**Table 1.** Categorization of COVID-19-associated susceptibility variants

|  | **Protective** | **Risk factor** | **Severity** | **Other** |
| --- | --- | --- | --- | --- |
| **CAV-FE** | 16 | 42 | 137 | 1 |
| **CAV** | 38 | 87 | 371 | 4 |
| **FCP + IP** | 37 | 29 | 4 | 70 |
| **FCP** | 3 | 6 | 16 | 0 |
| **IP** | 15 | 6 | 6 | 0 |
| **Total** | 109 | 170 | 534 | 75 |

Variant categories were generated based on the presumed effect and evidence level (Methods).

### Machine learning classifier and genome-wide prediction of novel COVID-19 variants

Variants in the category with the highest level of confidence, CAV-FE, were used to generate a machine learning classifier for severe COVID-19 variants. We selected a set of putatively neutral variants from gnomAD v2.1^13^ and a set of known disease-causing pathological mutations from the Human Gene Mutation Database (HGMD^®^)^14^ for training the classifier (Methods). To avoid selecting pathogenic variants that might also be associated with COVID-19 severity, non-COVID-19 pathogenic variants with Human Phenotype Ontology (HPO)^15^ terms related to abnormalities of the immune system, respiratory system, systemic blood pressure, or increased weight and diabetes, were excluded. Variants were annotated with 422 features at the variant, protein, gene and network level (Supplementary Table 1). The dataset was split into training and testing sets comprising 90 and 10 percent of the data respectively, and stratified by gene, such that genes represented by variants in the training set were not represented by variants in the testing set. We performed model selection and hyperparameter tuning using nested cross-validation with 5-fold outer and 5-fold inner validation loops with folds stratified by gene. RandomForest^16^ achieved the highest accuracy and Matthew’s correlation coefficient with the lowest variance across outer folds, and was selected as the final model (Extended Data Fig. 1). We subsequently performed hyperparameter tuning with RandomForest on the entire training dataset with 5-fold cross-validation for 500 iterations. The final RandomForest model successfully learned to discriminate between neutral/benign, non-COVID-19 pathogenic, and CAV-FE variants, achieving a macro-averaged ROC area under the curve (AUC) of 0.78 and an average precision (AP) of 0.74 (Fig. 2a,b). We further annotated all possible missense variants in the human genome (*n* = 82,468,698) and the COVID-19 severity computed predictions for these variants are available at https://itanlab.shinyapps.io/COVID19webpage/.

**Fig. 2:**
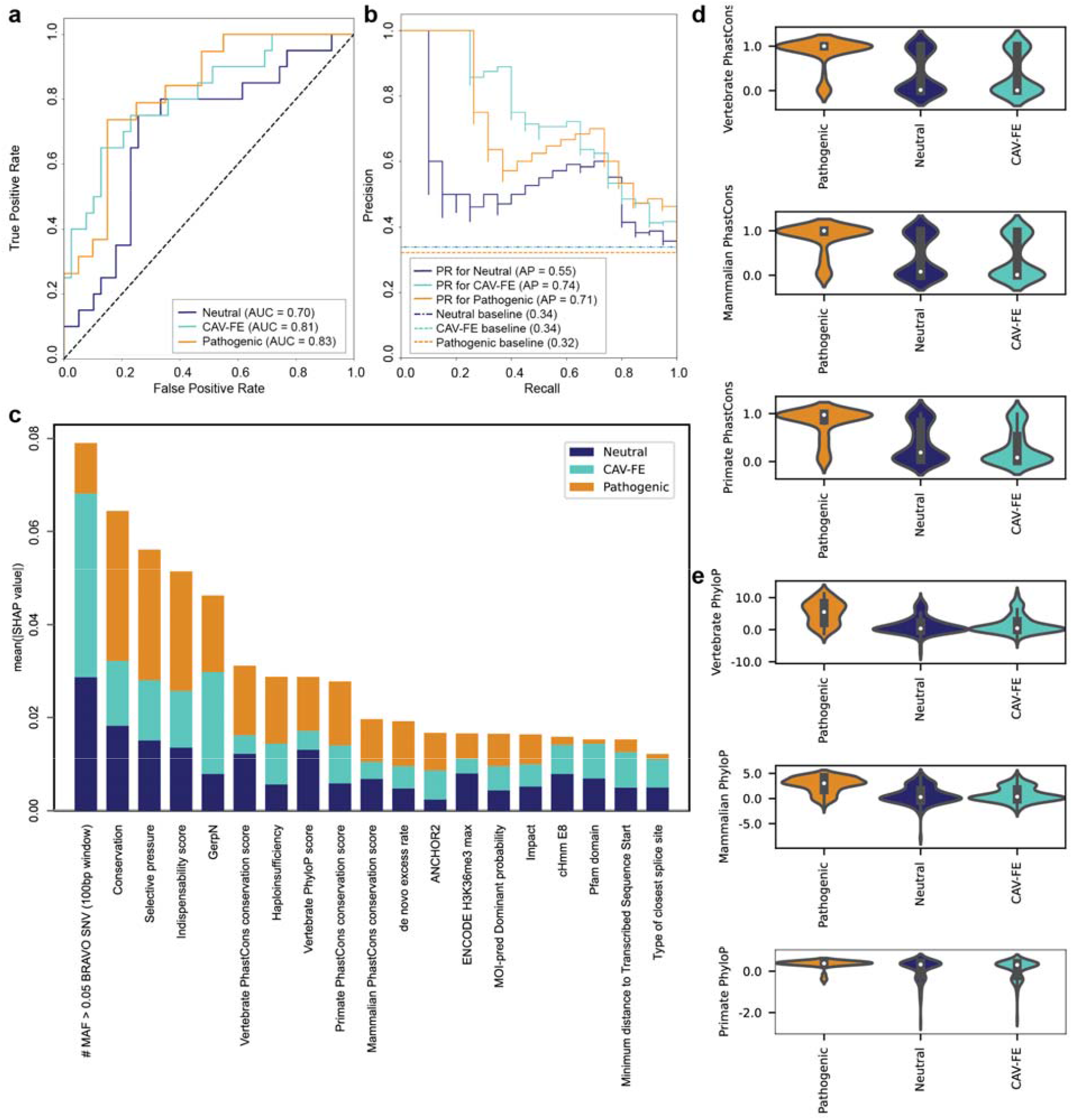
Evaluation of severe COVID-19 variant classifier and feature importance. **a**, ROC curves and **b**, precision-recall (PR) curves displaying the performance of the classification for neutral, non-COVID pathogenic and CAV-FE variants. AUC and AP values are shown in the legend. **c**, Bar charts demonstrating the mean SHAP values of the neutral, non-COVID pathogenic and CAV-FE variants for the features with the highest impact on the model. **d**, PhastCons and **e**, PhyloP scores of the three groups of variants. Violin plots display the median (white dot), interquartile range (IQR, thick gray bar in the center), and 25th percentile * 1.5 inter IQR and 75th percentile * 1.5 IQR (whiskers).

### Evaluation of feature importance

We calculated Shapley-value-based explanations^17^ (SHAP), a unified framework for the evaluation of feature importance, to investigate the contributions of individual features to the model’s predictions. SHAP revealed that the number of frequent (minor allele frequency, MAF > 0.05) BRAVO^18^ single nucleotide variants (SNV) in the 100 bp window surrounding a variant, variant conservation scores, and gene-level evolutionary pressure to be among the most impactful features of the model (Fig. 2c). Of the top twenty features identified by SHAP, six relate to the relative conservation of the variant site (Fig. 2c). Notably, we found that CAV-FE were consistently predicted to occur at significantly less conserved sites than the non-COVID-19 pathogenic variants and, in the case of the Primate PhastCons score^19^, significantly less conserved sites than neutral variants as well (Fig. 2d, Supplementary Table 2). In similar vein, CAV-FE were significantly more likely to have a higher number of frequent (MAF > 0.05) BRAVO variants occurring within 100 and 1000 base-pair windows than both non-COVID-19 pathogenic and neutral variants (Fig. 2e, Supplementary Table 3). Other impactful features included predictions of disordered protein binding residues^20^, the *de novo* mutation excess rate^21^, the indispensability score (which estimates gene essentiality based on a gene’s network and its evolutionary properties^22^), the maximum ENCODE H3K36me3 level from 10 cell lines^23^, gene haploinsufficiency^24^, among others (Fig. 2c). We also assessed these features in the three groups of variants using Benjamini-Hochberg (B-H)-adjusted pairwise *t*-tests. Disordered residues in protein structure provide high conformational flexibility, which enables binding to diverse interaction partners. These residues contribute to the activation of signaling processes through their context-dependent folding ability^25^. Compared to the non-COVID-19 pathogenic variants and neutral variants, CAV-FE variants were less likely to be associated with a disordered segment (*p* = 6 × 10^−6^ and 0.049, respectively). Consistent with their low level of conservation, CAV-FE displayed a lower *de novo* mutation excess rate and indispensability score than non-COVID-19 pathogenic variants (*p* = 3.6 × 10^−16^ and 1.57 × 10^−13^, respectively) whereas there was no statistically significant difference between CAV-FE and neutral variants in relation to these features (*p* = 0.248 and 0.201). Histone H3 trimethylation at lysine 36 is associated with actively transcribed regions and also plays an important role in heterochromatin composition^26^. CAV-FE had lower H3K36me3 levels than the neutral variants (*p* = 0.009) whereas no statistically significant difference was detected between non-COVID-19 pathogenic and CAV-FE variants (*p* = 0.056). Lastly, the haploinsufficiency score is a measure of the ability/inability of a gene carrying a single copy of a loss of function mutation to continue to function properly^24^. Genes carrying CAV-FE variants were more tolerant of haploinsufficiency as compared to those with non-COVID-19 pathogenic variants (*p* = 8.5 × 10^−5^) whereas the scores of CAV-FE and neutral variants were similar (*p* = 0.212).

### Gene-set and pathway enrichment analyses

We then performed gene enrichment and pathway analyses to gain further insight into the biological functions of the 68 genes harboring CAV-FE variants (Supplementary Table 4). InnateDB pathway analysis^27^ revealed 117 significantly over-represented pathways (B-H-adjusted *p* < 0.05), the majority of which function in innate and adaptive immune responses (Supplementary Table 5). The most significant pathways from the KEGG database^28^ were toll-like receptor signaling pathway (*p* = 2.06 × 10^−10^), hepatitis C (*p* = 3.62 × 10^−10^), cytokine-cytokine receptor interaction (*p* = 1.34 × 10^−6^) and natural killer cell-mediated cytotoxicity (*p* = 4.20 × 10^−5^) whilst interferon alpha/beta signaling (*p* = 6.22 × 10^−10^), toll-like receptor cascades (*p* = 6.48 × 10^−8^), activation of IRF3/IRF7 mediated by TBK1/IKK epsilon (*p* = 6.11 × 10^−7^) and activated TLR4 signaling (*p* = 1.72 × 10^−6^) were the most significantly over-represented pathways from Reactome^29^ (Fig. 3a). The canonical pathway analyses using Ingenuity Pathway Analysis (IPA) software (QIAGEN Inc., http://www.qiagen.com/ingenuity) supported the results of InnateDB by returning 88 significantly enriched pathways, most of which were related to the immune system and infection (Supplementary Table 6). Further, the results of IPA revealed that hypercytokinemia/hyperchemokinemia in the pathogenesis of influenza (*p* = 1.99 × 10^−19^), the coronavirus pathogenesis pathway (*p* = 6.31 × 10^−19^), the neuroinflammation signaling pathway (*p* = 3.09 × 10^−10^) and the pathogen-induced cytokine storm signaling pathway (*p* = 8.32 × 10^−10^) were the most significant pathways (Fig. 3a). We then evaluated significantly enriched GO terms in our high-confidence COVID-19 gene set using InnateDB GO analysis. The top results included defense response to virus (*p* = 6.33 × 10^−13^), type I interferon signaling pathway (*p* = 2.21 × 10^−12^), innate immune response (*p* = 3.43 × 10^−10^), response to virus (*p* = 3.30 × 10^−10^) and cytokine-mediated signaling pathway (*p* = 3.48 × 10^−9^) (Extended Data Fig. 2, Supplementary Table 7).

**Fig. 3:**
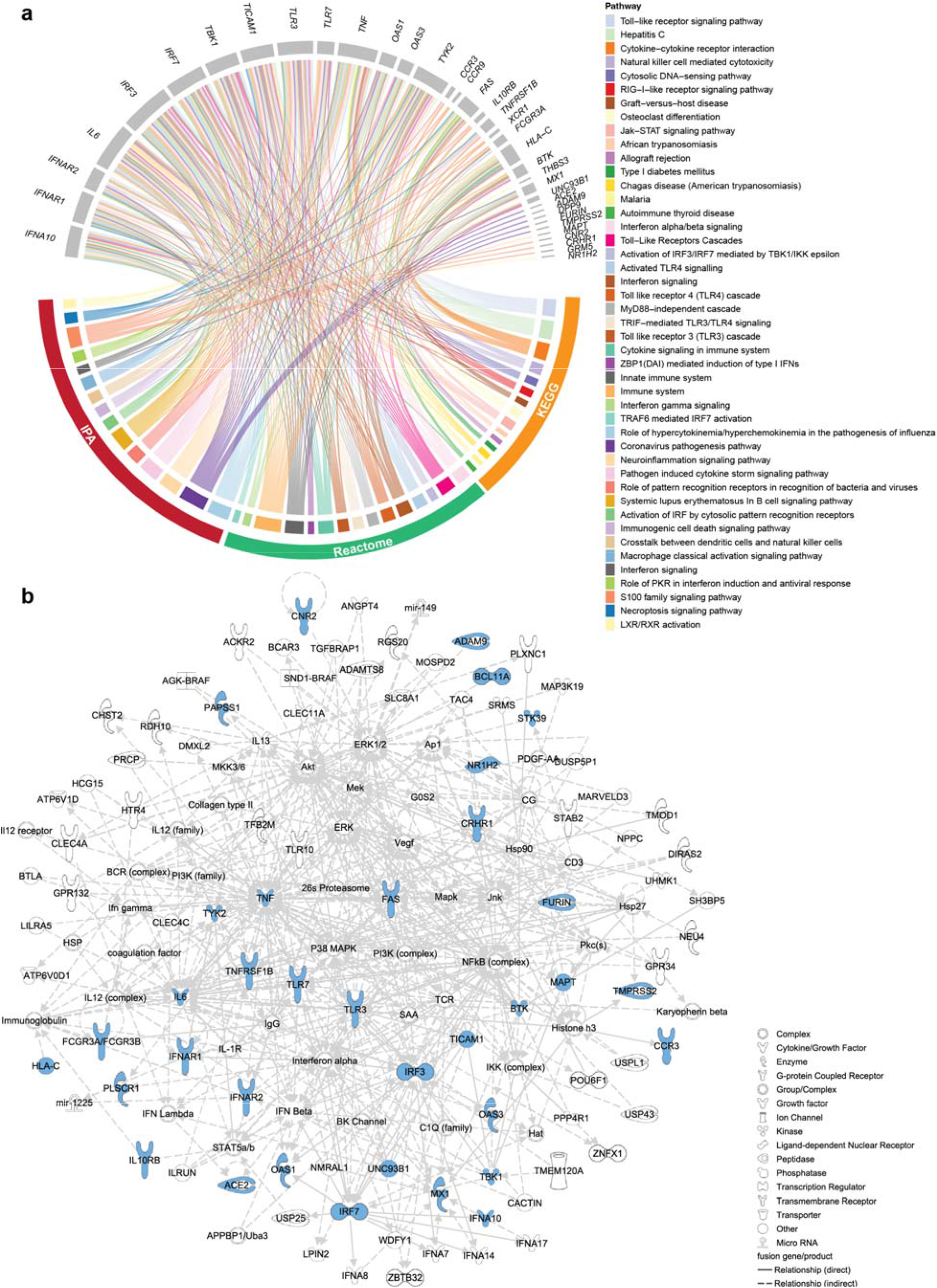
Pathway and IPA network analyses. **a**, Circos plot showing the top 15 pathways identified by InnateDB (KEGG and Reactome) and IPA. The order of pathways on the plot is according to their *p* values from most to least significant in each pathway database. **b**, Subnetwork associated with “microbial response, cellular development and inflammatory response” from IPA network analysis.

Next, we explored biological networks to obtain a deeper understanding of the interactions between the high-confidence COVID-19 genes and biomolecules. IPA network analysis returned a network with a *p* value of 10^−57^, which included 35 genes harboring CAV-FE variants (Fig. 3b). Interestingly, the top diseases and functions associated with this network were antimicrobial response, cellular development and inflammatory response (Supplementary Table 8). We also used the NetworkAnalyst tool to investigate protein-protein interaction (PPI) networks within high-confidence COVID-19 genes (Supplementary Table 9). The subnetwork that was generated based on the STRING interactome^30^ included 25 genes all of which were also identified by the IPA network analysis (Extended Data Fig. 3). The difference between the two networks likely arose from the involvement of both direct and indirect relationships in the IPA network analysis as opposed to STRING, which uses only direct PPIs. We then analyzed the significantly enriched diseases and biological functions in the high-confidence COVID-19 gene set. The results obtained indicated viral infections including COVID-19 and antiviral response, and also inflammatory and auto-inflammatory diseases such as systemic lupus erythematosus (SLE), inflammatory demyelinating disease and multiple sclerosis (Fig. 4a, Supplementary Table 10). Consistent with the results of IPA, Enrichr HPO analysis revealed recurrent viral infections (HP:0004429, *p* = 4.21 × 10^−4^) as the most significantly enriched HPO term^15,31^ (Supplementary Table 11).

**Fig. 4:**
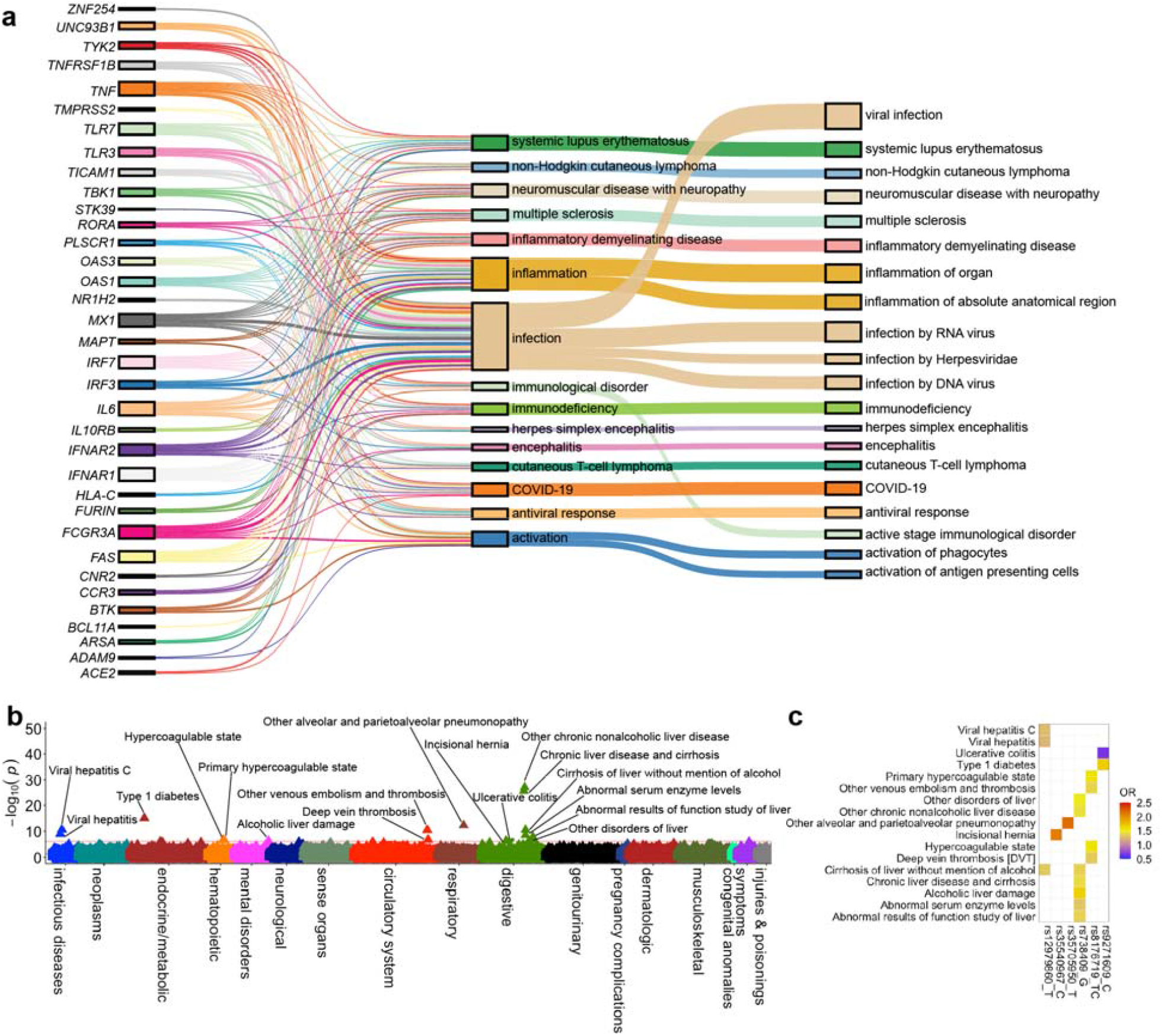
IPA diseases/functions and PheWAS in the BioMe BioBank. **a**, Top 20 diseases/functions associated with the genes harboring CAV-FE variants by IPA. Boxes on the left depict the genes, boxes in the middle represent their function, whereas the boxes on the right denote the disease/function annotations. **b**, Results of transethnic meta-PheWAS in individuals with AA, EA and HA ancestries (*n* = 39,386) using 288 clumped COVID-19 variants. The red line denotes the phenome-wide significance threshold calculated by means of the B-H procedure using an FDR of 0.01. Phenotypes significantly associated with COVID-19 variants are labeled. The direction of triangles represents the direction of effect. **c**, Heatmap displaying ORs of the significant associations. The x axis shows the variants and effect alleles used in PheWAS; the y axis displays various phenotype descriptions.

### PheWAS in the Mount Sinai BioMe BioBank

We performed PheWAS using genotypes and electronic health records (EHR) of 39,386 study participants with African American (AA), European American (EA) and Hispanic American (HA) ancestries from the Mount Sinai BioMe BioBank. We clustered known COVID-19 variants according to linkage disequilibrium (LD) in order to obtain a list of independent variants associated with COVID-19 (Methods). The number of independent variants based on LD was 285 in AA, 286 in EA and 288 in HA cohorts across 458, 466 and 629 phenotypes, respectively. PheWAS in AA and EA groups revealed a significant association of the rs9271609-C allele at the *HLA-DQA1* locus with type 1 diabetes mellitus (DM) (Extended Data Fig. 4a, b and Supplementary Tables 12 and 13). Associations of rs738409-G in *PNPLA3* with chronic nonalcoholic liver disease and cirrhosis were noted in PheWAS of individuals with EA and HA ancestry (Extended Data Fig. 4b, c and Supplementary Tables 13 and 14). Moreover, rs738409-G was significantly associated with abnormal results of liver function studies and the detection of abnormal serum enzyme levels in the PheWAS of the HA cohort. The other significant finding from PheWAS in the HA cohort was the association of rs35705950 in the proximity of *MUC5B* with alveolar or parietoalveolar pneumonitis. Our findings support the results of previous studies that showed the rs35705950-T allele being associated with idiopathic pulmonary fibrosis while being a protective variant for COVID-19^32^. Finally, PheWAS in the HA cohort also revealed an association of the rs12979860-T allele in *IFNL4* with viral hepatitis and cirrhosis. The results of the transethnic meta-analysis highlighted another well-known genetic association, the association of the ABO locus (rs8176719-TC) with a hypercoagulable state and deep vein thrombosis^33^. The remainder of the meta-analysis results yielded comparable findings to the ancestry-specific PheWAS (Fig. 4b, Supplementary Table 15).

### The web-based interface of the COVID-19 Host Genetic Variants

Finally, we generated a web-based interface allowing users to search and download published and predicted COVID-19-associated variants, which are publicly available at https://itanlab.shinyapps.io/COVID19webpage/ (Fig. 1). The website contains genomic positions based on genome assembly hg38, Ensembl annotations and HGVS nomenclature for the 820 known COVID-19 susceptibility host genetic variants, as well as their effects on COVID-19 phenotype, accession numbers of the respective studies and assigned categories based on the confidence level. Further, users can submit additional host genetic variants associated with COVID-19 susceptibility using the submission form on the website. We have also made accessible the PheWAS results for the known COVID-19 variants on a separate tab. The precomputed COVID-19 predictions for all possible missense variants are also provided on the website for exploration and downloading. The web server will be updated by regularly screening new publications and user submissions.

## Discussion

Identification of the genetic risk factors that contribute to human disease is an essential component of precision medicine and genomics. These data-driven approaches require the consolidation of disparate data sources and leveraging the data by developing proper analytical methods for the discovery of novel findings. By systematic evaluation of the host genetic variants involved in COVID-19 susceptibility, we determined 820 variants, of which 196 belonged to the category with the highest confidence level. The majority of the variants (91.33%) in the CAV-FE category have been suggested to be risk factors or associated with severe COVID-19 whereas the rest have been reported as protective variants. The machine learning classifier that we developed using CAV-FE variants in the severity and risk factor categories successfully distinguished CAV-FE variants from non-COVID-19 pathogenic and neutral variants. The model enabled us to estimate the prediction scores for all possible human missense variants. The assessment of the variant-level features that contributed to the model set forth lower conservation levels of CAV-FE variants compared to non-COVID-19 pathogenic and neutral variants. It is important to note that the degree of conservation of a sequence does not always correlate with its functionality, which can be exemplified by the positive selection events elevating the frequency of functional variants^34^. Therefore, further investigation of the variant constraints is required to understand if COVID-19-associated variants are subject to evolutionary pressure. With respect to the gene-level interpretation, we expected to obtain comparable scores with CAV-FE and neutral variants for both the *de novo* mutation excess rate and indispensability score, since we selected neutral variants from the same set of genes that CAV-FE occurred in (Methods).

Pathway and GO enrichment analyses in genes harboring CAV-FE variants revealed interferon signaling, toll-like receptor pathway and cytokine-mediated signaling, all of which were implicated in the pathogenesis of severe COVID-19^1,4-6^. The most significant network identified by IPA contained about half of the genes carrying CAV-FE variants. The molecules identified in the network were found to be associated with antimicrobial response, cellular development, and inflammatory response consistent with their biological functions. Interactions between the focus molecules and the other components of the network, such as the coagulation factor, support the previous studies on the pathogenesis of severe COVID-19^35^. Therefore, the rest of the molecules in this network can be prioritized as candidates for future investigations in the pathogenesis of COVID-19 and for the discovery of druggable targets. Similarly, the significantly enriched diseases and functions included COVID-19, several other viral infections and immunodeficiency, as well as autoimmune diseases including SLE. These results support the previous studies suggesting that *TYK2* has opposing associations with COVID-19 susceptibility and predisposition to autoimmune conditions^32,36^. Notably, the results of all gene-set enrichment and pathway analyses are indicative of a role for COVID-19-associated genes in essential immune processes, thereby emphasizing the likely importance of underlying genetic variation on the impaired host immune response to COVID-19^1,3^.

The availability of a large set of COVID-19-associated variants provided us with an unparalleled opportunity to examine the genetic factors shared between COVID-19 susceptibility/severity and comorbidities that might represent risk factors for worse outcomes in COVID-19 disease. PheWAS of individuals with AA and EA ancestries revealed the association of the rs9271609-T allele in *HLA-DQA1* with Type 1 DM. The rs9271609-T allele has been implicated in COVID-19 severity^35^. Polymorphisms in class II HLA genes, *HLA-DQ* and *HLA-DR*, are well-established determinants for Type 1 DM susceptibility^37^. Although previous reports have proposed an increased risk of severe COVID-19 for patients with Type 1 DM, the opposite association noted in the current results provides some support for those studies that have argued for a relationship between autoimmune conditions and a reduced risk of severe COVID-19^32,36^. The ancestry-specific PheWAS results reflect the well-validated association of the rs738409-G allele with various types of chronic liver disease^38^, which is known to be especially prominent in the Hispanic population^39^. Individuals with established liver disease have been shown to have an increased risk of hospitalization due to COVID-19 in a previous study^40^, although there is no consensus as to the effect of chronic liver disease and cirrhosis on COVID-19 severity^40^. There are nevertheless conflicting interpretations regarding the impact of rs738409 on the course of COVID-19. GG genotype for rs738409 has been shown to increase the risk of severe disease in patients younger than 65 years^41^ whilst the rs738409-G allele has been reported to be associated with a reduced risk of hospitalization and mortality in another investigation^42^. Also, the association of the rs12979860-T allele in *IFNL4* with viral hepatitis and cirrhosis was significant in the HA group. Carriers of the rs12979860-T allele have been previously shown to experience persistent hepatitis C virus (HCV) infection due to a weaker host immune response against HCV^43,44^. In addition, the minor allele, rs12979860-T, has been proposed as a risk factor for severe COVID-19^45^ whereas the CC genotype for the major allele, rs12979860-C, has been reported to occur with a higher incidence in COVID-19 patients compared to healthy controls^46^.

Hypercoagulable state and deep vein thrombosis were found to be associated with rs8176719 in the transethnic meta-PheWAS. The role of genetic variants at the ABO locus in COVID-19 susceptibility and severity is well-established^47,48^. The prevalence of severe disease is higher in patients with blood group A whereas blood group O is associated with a lower risk for severe COVID-19^48^. Following a similar approach to that of the current study, two previous studies on the Million Veterans Program and UK BioBank datasets respectively, evaluated the pleiotropic effects of a set of severe COVID-19-associated variants^32,36^. Both studies performed their analyses using a larger cohort and identified a number of significant phenotypes, which included pre-existing conditions that increase the risk of having severe illness from COVID-19, such as thromboembolism, type 2 DM and hypercholesterolemia. Overall, our PheWAS results support previously reported disease associations, but also revealed new associations pertaining to a shared genetic etiology between COVID-19 susceptibility and various other pre-existing conditions.

In summary, this study provides the scientific community with a knowledgebase of the known genetic determinants of COVID-19 susceptibility and severity, along with biological insights into immune system-related functions of genes implicated in COVID-19 pathogenesis and phenotype associations revealing the pleiotropic effects of COVID-19 susceptibility variants on underlying risk factors for COVID-19 severity. The categorization of a comprehensive set of previously published variants based on evidence level will enable rapid evaluation of these variants by researchers. By employing the first available machine learning classifier for the prediction of COVID-19 host genetic variants, our work also provides genome-wide predictions for all possible human missense variants which should facilitate the discovery of novel genetic risk factors for severe COVID-19.

## Methods

### Curation and categorization of COVID-19-associated variants

The literature search in PubMed was conducted in order to identify studies that have investigated host genetic variants associated with COVID-19 susceptibility/severity. The combinations ‘COVID19,’ ‘coronavirus’, ‘SARS’ and ‘COVID-19’ with ‘genetics’, ‘GWAS’, ‘genetic factor’, ‘genetic susceptibility’, ‘genetic association’, ‘genetic resistance’, ‘genetic variant’, ‘genetic variation’, ‘genotype’, ‘host factor’ or ‘human genome’ were used as search terms. The search for publications up until August 5, 2022 yielded 1,977 records. 222 publications remained after the review of titles and abstracts led to the exclusion of studies that did not contain data pertaining to COVID-19-associated host genetic variants. The second round of review comprised the careful manual curation of variants, as well as the gathering of information about study type, analytical methods and study population from the main text and supplementary material of publications. The final list of 820 COVID-19-associated variants were converted to genomic positions based on human genome assembly hg38. The effect of variants on disease phenotype was ascertained as ‘Risk factor’, ‘Severity’, ‘Protective’ and ‘Other’ according to the findings in the respective studies. Variants were further categorized into five levels of confidence based on the published evidence from the highest level of confidence to the lowest as follows: CAV-FE, CAV, FCP + IP, FCP and IP. Where multiple studies identified the same variant, the combined level of evidence was used for categorization of the variant.

### Machine learning classifier

To develop the classifier of COVID-19 severity-associated variants, we collected putatively neutral variants from gnomAD v2.1 after removing those variants categorized as disease-causing mutations (DM) in the Human Gene Mutation Database (HGMD) 2022.2 Professional release, making the assumption that the majority of the remaining gnomAD variants are benign. Additionally, we selected a set of non-COVID-19 pathogenic variants from the DM category of the HGMD. To avoid selecting pathogenic variants that might also be associated with COVID-19 severity, we filtered the HGMD dataset such that all variants resulting in HPO^15^ phenotypes related to abnormalities of the immune system (HP:0002715), respiratory system (HP:0002086), systemic blood pressure (HP:0030972), increased weight (HP:0004324), and diabetes (HP:0000819) were removed. 134 non-COVID-19 pathogenic variants were then randomly selected from the genes in which the CAV-FE occurred, matching the number of CAV-FE per gene wherever possible. An additional 23 non-COVID-19 pathogenic regulatory variants were selected to match the 23 non-genic CAV-FE. 134 gnomAD variants were similarly randomly selected from the genes in which CAV-FE variants occurred matching the number of CAV-FE per gene where possible. 23 non-genic gnomAD variants were selected to match the 23 non-genic CAV-FE. The full dataset was split such that variant genes did not overlap between the training and testing sets. The training set consisted of 138 non-COVID pathogenic variants, 137 CAV-FE, and 137 neutral variants, whilst the testing set comprised 19 non-COVID pathogenic variants, 20 CAV-FE and 20 neutral variants.

To estimate the performance and variance associated with our data preprocessing, model architecture and model hyperparameter selection pipeline, we employed 5-fold outer, 5-fold inner nested cross-validation. Folds were stratified by variant gene such that training and testing folds did not contain variants from the same genes. Model performance was assessed by means of the Matthew’s correlation coefficient, macro averaged F1-score, and accuracy. We then applied 5-fold cross-validation on all the training data with the best model to determine the final preprocessing and model hyperparameters.

#### Feature importance

SHAP values were generated for the model’s predictions for all variants in the testing set with the SHAP python library’s TreeExplainer using default parameters. For the statistical analysis of features, we performed two-sample, one-sided *t*-tests comparing the mean of the CAV-FE variants to the mean of the non-COVID pathogenic and benign variants respectively. We employed B-H correction to control the false discovery rate at 0.05 (Supplementary Tables 2, 3).

### Enrichment analyses

68 genes harbored CAV-FE variants included in the enrichment analyses (Supplementary Table 4). Enrichment of the genes in pathways and GO terms was investigated with InnateDB version 5.4^27^. Major pathway resources including KEGG^28^, Reactome^29^, NetPath^49^, INOH^50^ and PID^51^ are incorporated in the InnateDB pathway analysis for the exploration of over-represented biological pathways in a particular gene or protein set. Pathway enrichment analysis, network analysis and enrichment analysis of biological functions and diseases in the high-confidence COVID-19 gene set were performed using IPA software version 01-21-03 (http://www.qiagen.com/ingenuity) with default parameters except for selecting the number of molecules per network as 140 and restricting the analysis to ‘human’. The subnetworks of PPIs were evaluated using the NetworkAnalyst^52^ tool on the high-confidence STRING^30^ interactome with medium (400) to high (1000) confidence score and a minimum network option for displaying key connectivities. Enrichr^31^ web server was used for the gene set enrichment analysis of HPO^15^ terms. FDR adjustment was performed using B-H method for the *p* values in all analyses.

### PheWAS

#### Cohort

The Mount Sinai BioMe BioBank contains array genotyping data from 53,982 individuals who were admitted to Mount Sinai primary care clinics. Since no phenotype-specific selection criteria were applied during the recruitment of participants, the BioMe BioBank may be considered to be a representative subset of the general population of New York City and its environs. The three major ancestries in the BioMe BioBank were AA (*n* = 9,616), EA (*n* = 13,401) and HA (*n* = 16,369), which were included in the downstream analyses.

#### Genotype data quality control (QC)

The Mount Sinai BioMe BioBank comprises two different batches of array genotyping data that were produced using the Infinium Global Screening Array (GSA, 31,683 samples and 635,623 variants) and the Infinium Global Diversity Array (GDA, 22,299 samples and 1,833,111 variants). First, individuals were stratified by ancestry. Then, duplicated samples (which were detected using KING^53^), samples with sex discrepancies, samples with a call rate < 95% or a heterozygosity rate that fell outside of three standard deviations from the mean, were removed from each ancestral group. Variants with a call rate < 95% and a Hardy-Weinberg equilibrium (HWE) *p*-value < 1e-8 were filtered out. We obtained a final combined dataset of 53,449 individuals and 335,682 variant sites, which were imputed using TOPMed^18^. The imputed dataset contained 60,491,206 variants with an imputation accuracy, *R*^*2*^ > 0.7.

#### PCA

Variants with a HWE *p*-value < 5e-6 were filtered out and were pruned according to linkage disequilibrium (LD) using an *r*^*2*^ threshold of 0.5. Variants with a minor allele frequency ≥ 1% were selected. PCA was performed with Plink v.1.9^54^.

#### Ancestry-specific and meta-PheWAS

COVID-19-associated host genetic variants were clumped together according to LD patterns. Variants were clustered using an *r*^*2*^ threshold of 0.1 and 250 kb window size in the AA, EA and HA samples of the BioMe BioBank. Three ancestry-specific PheWAS were performed using 285, 286 and 288 clustered variants in AA, EA and HA groups, respectively, for testing their associations with phenotypes extracted from the EHR data. ICD10-CM codes of the participants were mapped to phecodes using the R PheWAS package^55^. Phenotypes with at least 100 cases were retained, resulting in 458, 466 and 629 tested phenotypes for AA, EA and HIS ancestries, respectively. Age, biological sex, sequencing batch and first 20 PCs were included in the analyses as covariates. A trans-ethnic meta-analysis was performed using phewasMeta function and the results of PheWAS in three groups. B-H procedure was followed in order to control the false discovery rate (FDR) at 0.01.

### The web-based interface for COVID-19 Host Genetic Variants

The R-Shiny platform (https://CRAN.R-project.org/package=shiny) was used to generate the web-based interface for COVID-19 Host Genetic Variants.

## Data Availability

All data produced in the present work are contained in the manuscript and are available online at https://itanlab.shinyapps.io/COVID19webpage/

## Data availability

The web-based interface for COVID-19 host genetic variants can be accessed publicly at https://itanlab.shinyapps.io/COVID19webpage/ and may be used for all non-commercial purposes.

## Acknowledgements

This work was supported by The Charles Bronfman Institute for Personalized Medicine. P.D.S. and D.N.C. acknowledge the financial support of Qiagen Inc. through a License Agreement with Cardiff University.

---

**Extended Data Fig. 1:**
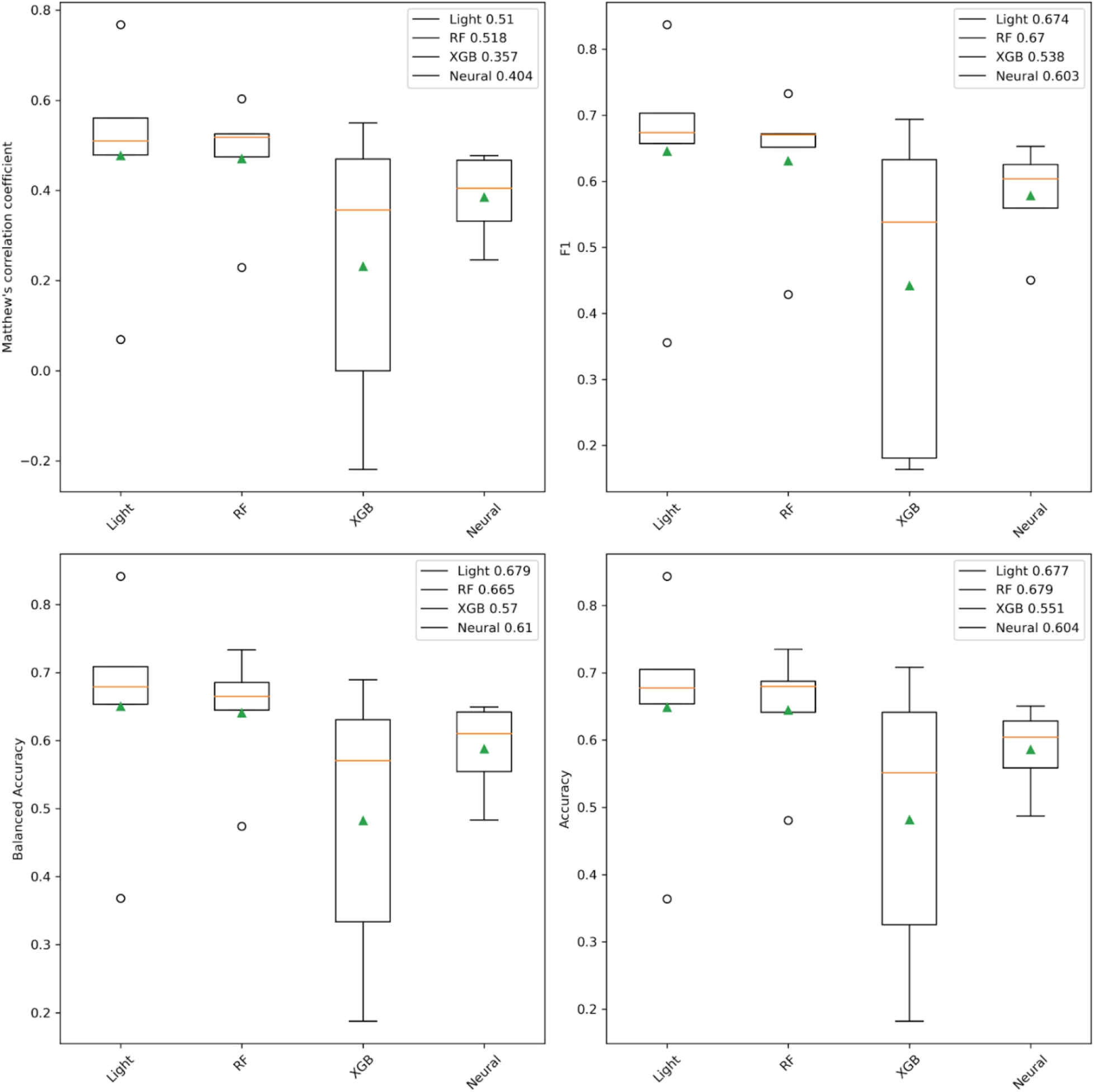
Model selection. Box plots displaying the measures of performance assessment for the LightGBM (Light), RandomForest (RF), XGBoost (XGB) and Neural Network (Neural) algorithms.

**Extended Data Fig. 2:**
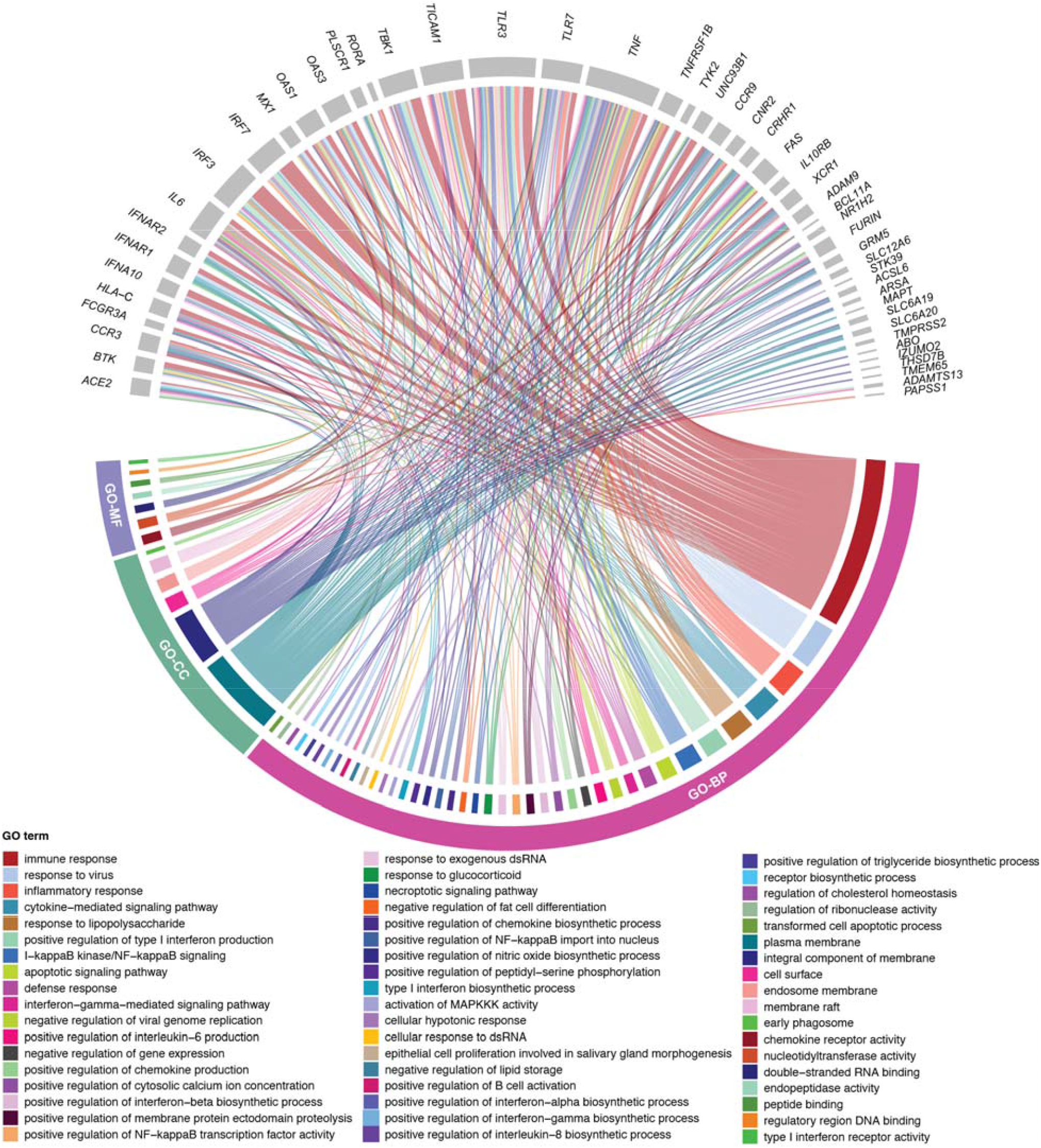
InnateDB GO analysis. Circos plot depicting the most significantly over-represented GO terms (B-H-adjusted *p* < 0.01) in the high-confidence COVID-19 gene set. The child terms were combined under their parents, if the parent term was also in the list of top terms. BP: biological process, CC: cellular component, MF: molecular function.

**Extended Data Fig. 3:**
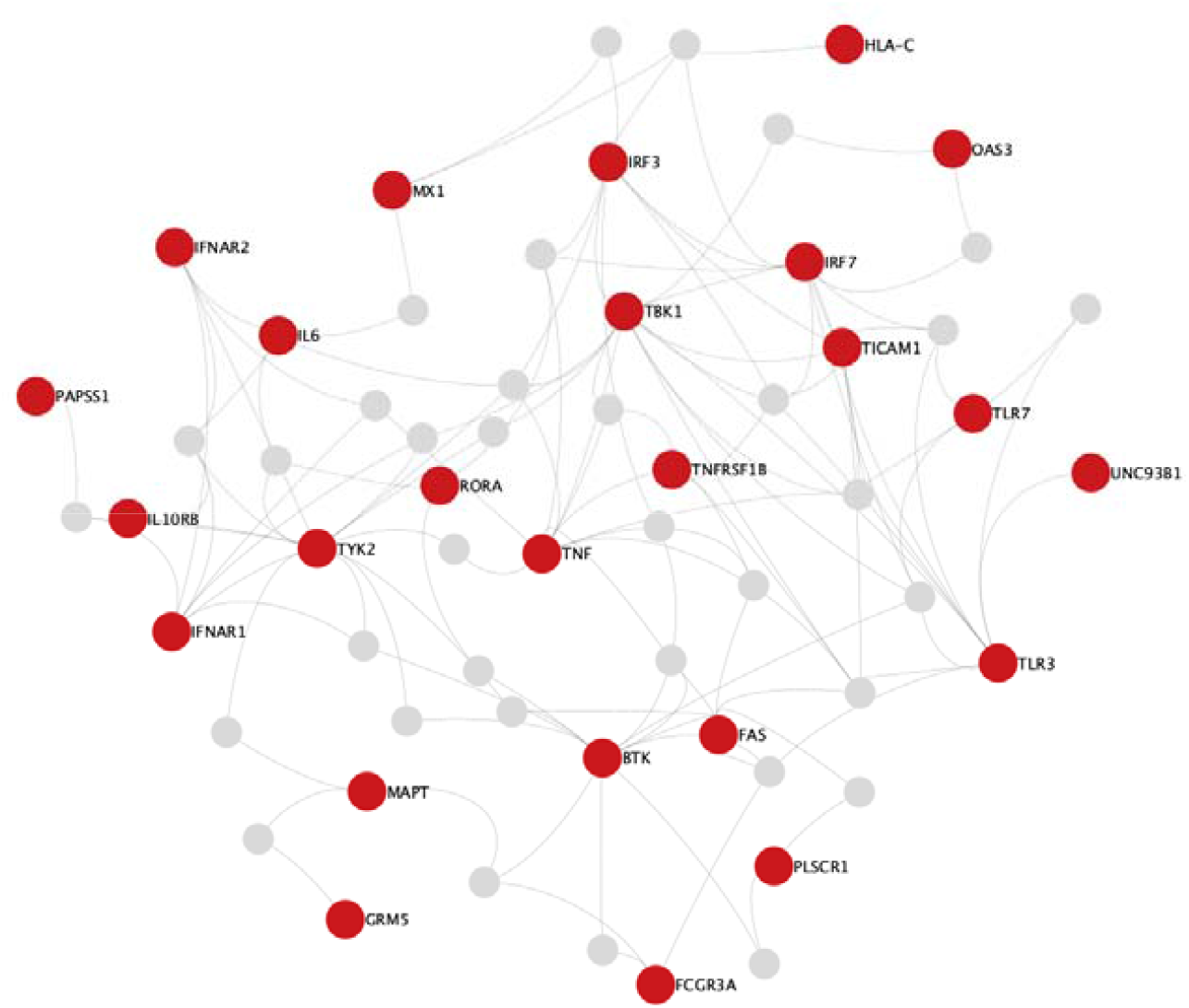
STRING PPI subnetwork identified by the NetworkAnalyst tool. The red nodes display genes carrying CAV-FE variants.

**Extended Data Fig. 4:**
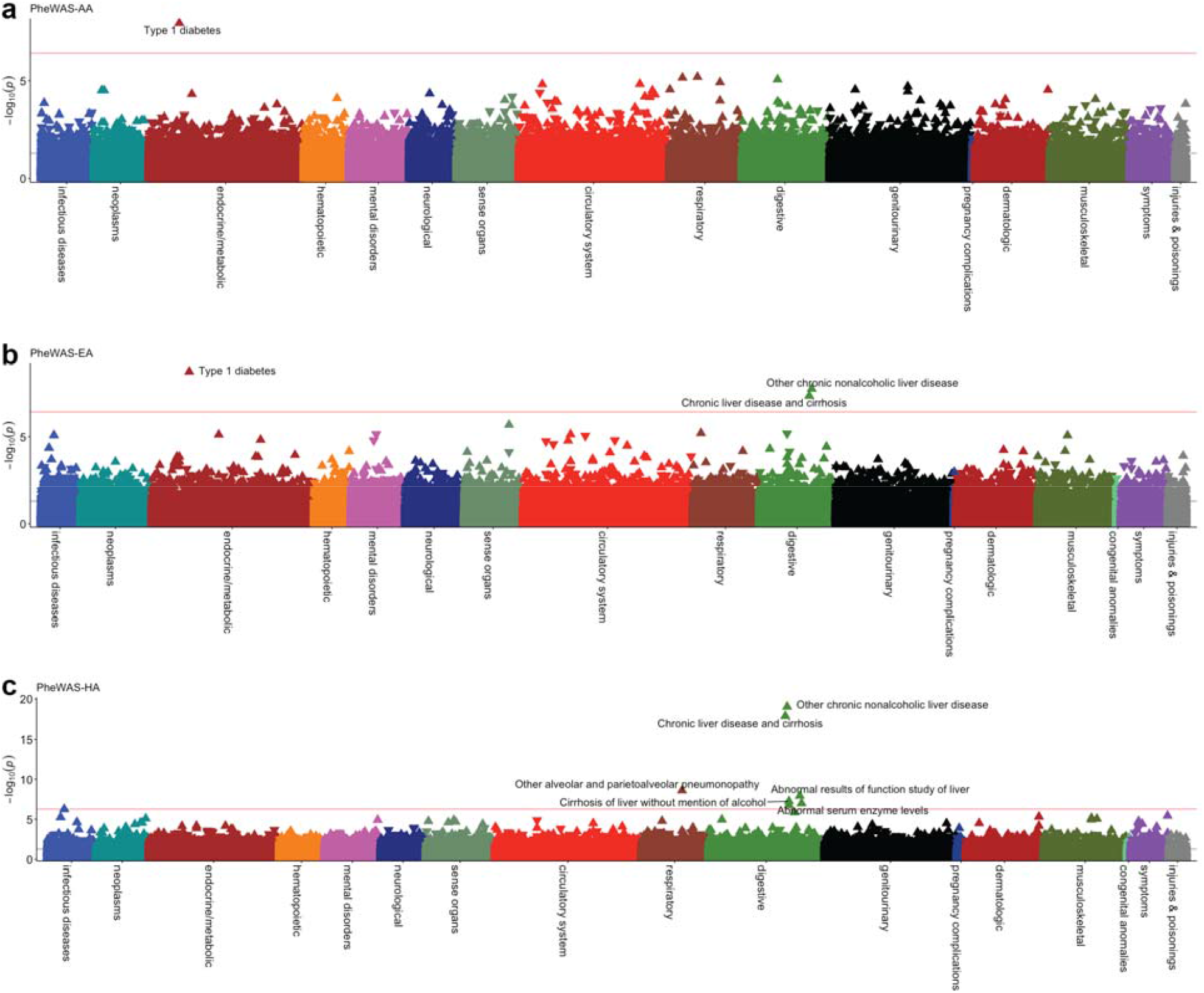
Ancestry-specific PheWAS. Results of PheWAS in **a**, the AA ancestry (*n* = 9,616) using 285 clumped COVID-19 variants. **b**, the EA ancestry (*n* = 13,401) using 286 clumped COVID-19 variants **c**, the HA ancestry (*n* = 16,369) using 288 clumped COVID-19 variants. Red lines denote the phenome-wide significance thresholds calculated by means of the B-H procedure using an FDR of 0.01. Phenotypes significantly associated with COVID-19 variants are labeled. The direction of triangles represents the direction of effect.

## References

1. Zhang, Q. et al. Inborn errors of type I IFN immunity in patients with life-threatening COVID-19. Science 370, eabd4570 (2020).

2. Severe Covid-19 GWAS Group. Genomewide association study of severe Covid-19 with respiratory failure. N. Engl. J. Med. 383, 1522–1534 (2020).

3. Pairo-Castineira, E. et al. Genetic mechanisms of critical illness in COVID-19. Nature 591, 92–98 (2021).

4. COVID-19 Host Genetics Initiative. Mapping the human genetic architecture of COVID-19. Nature 600, 472–477 (2021).

5. Asano, T. et al. X-linked recessive TLR7 deficiency in ∼1% of men under 60 years old with life-threatening COVID-19. Sci. Immunol. 6, eabl4348 (2021).

6. Zhang, Q. et al. Recessive inborn errors of type I IFN immunity in children with COVID-19 pneumonia. J. Exp. Med. 219, e20220131 (2022).

7. Wu, P. et al. Trans-ethnic genome-wide association study of severe COVID-19. Commun. Biol. 4, 1034 (2021).

8. Dite, G.S., Murphy, N.M. & Allman, R. An integrated clinical and genetic model for predicting risk of severe COVID-19: a population-based case-control study. PLoS One 16, e0247205 (2021).

9. Guo, X., Chen, Z., Xia, Y., Lin, W. & Li, H. Investigation of the genetic variation in ACE2 on the structural recognition by the novel coronavirus (SARS-CoV-2). J. Transl. Med. 18, 321 (2020).

10. Ji, X.S., Chen, B., Ze, B. & Zhou, W.H. Human genetic basis of severe or critical illness in COVID-19. Front. Cell Infect. Microbiol. 12, 963239 (2022).

11. Kwok, A.J., Mentzer, A. & Knight, J.C. Host genetics and infectious disease: new tools, insights and translational opportunities. Nat. Rev. Genet. 22, 137–153 (2021).

12. van der Made, C.I., Netea, M.G., van der Veerdonk, F.L. & Hoischen, A. Clinical implications of host genetic variation and susceptibility to severe or critical COVID-19. Genome Med. 14, 96 (2022).

13. Karczewski, K.J. et al. The mutational constraint spectrum quantified from variation in 141,456 humans. Nature 581, 434–443 (2020).

14. Stenson, P.D. et al. The Human Gene Mutation Database (HGMD^®^): optimizing its use in a clinical diagnostic or research setting. Hum. Genet. 139, 1197–1207 (2020).

15. Kohler, S. et al. The Human Phenotype Ontology in 2021. Nucleic Acids Res. 49, D1207–D1217 (2021).

16. Breiman, L. Random Forests. Machine Learning 45, 5–32 (2001).

17. Lundberg, S.M. et al. From local explanations to global understanding with explainable AI for trees. Nat. Mach. Intell. 2, 56–67 (2020).

18. Taliun, D. et al. Sequencing of 53,831 diverse genomes from the NHLBI TOPMed Program. Nature 590, 290–299 (2021).

19. Siepel, A. et al. Evolutionarily conserved elements in vertebrate, insect, worm, and yeast genomes. Genome Res. 15, 1034–1050 (2005).

20. Meszaros, B., Erdos, G. & Dosztanyi, Z. IUPred2A: context-dependent prediction of protein disorder as a function of redox state and protein binding. Nucleic Acids Res. 46, W329–W337 (2018).

21. Samocha, K.E. et al. A framework for the interpretation of de novo mutation in human disease. Nat. Genet. 46, 944–950 (2014).

22. Khurana, E., Fu, Y., Chen, J. & Gerstein, M. Interpretation of genomic variants using a unified biological network approach. PLoS Comput. Biol. 9, e1002886 (2013).

23. Rosenbloom, K.R. et al. ENCODE whole-genome data in the UCSC Genome Browser: update 2012. Nucleic Acids Res. 40, D912–917 (2012).

24. Huang, N., Lee, I., Marcotte, E.M. & Hurles, M.E. Characterising and predicting haploinsufficiency in the human genome. PLoS Genet. 6, e1001154 (2010).

25. van der Lee, R. et al. Classification of intrinsically disordered regions and proteins. Chem. Rev. 114, 6589–6631 (2014).

26. Chantalat, S. et al. Histone H3 trimethylation at lysine 36 is associated with constitutive and facultative heterochromatin. Genome Res. 21, 1426–1437 (2011).

27. Breuer, K. et al. InnateDB: systems biology of innate immunity and beyond--recent updates and continuing curation. Nucleic Acids Res. 41, D1228–1233 (2013).

28. Kanehisa, M., Furumichi, M., Tanabe, M., Sato, Y. & Morishima, K. KEGG: new perspectives on genomes, pathways, diseases and drugs. Nucleic Acids Res. 45, D353–D361 (2017).

29. Joshi-Tope, G. et al. Reactome: a knowledgebase of biological pathways. Nucleic Acids Res. 33, D428–432 (2005).

30. Szklarczyk, D. et al. STRING v10: protein-protein interaction networks, integrated over the tree of life. Nucleic Acids Res. 43, D447–452 (2015).

31. Kuleshov, M.V. et al. Enrichr: a comprehensive gene set enrichment analysis web server 2016 update. Nucleic Acids Res. 44, W90–97 (2016).

32. Verma, A. et al. A Phenome-wide association study of genes associated with COVID-19 severity reveals shared genetics with complex diseases in the Million Veteran Program. PLoS Genet. 18, e1010113 (2022).

33. Wu, O., Bayoumi, N., Vickers, M.A. & Clark, P. ABO(H) blood groups and vascular disease: a systematic review and meta-analysis. J. Thromb. Haemost. 6, 62–69 (2008).

34. Ponting, C.P. Biological function in the twilight zone of sequence conservation. BMC Biol. 15, 71 (2017).

35. Kousathanas, A. et al. Whole-genome sequencing reveals host factors underlying critical COVID-19. Nature 607, 97–103 (2022).

36. Regan, J.A. et al. Phenome-wide association study of severe COVID-19 genetic risk variants. J. Am. Heart Assoc. 11, e024004 (2022).

37. Atkinson, M.A. & Eisenbarth, G.S. Type 1 diabetes: new perspectives on disease pathogenesis and treatment. Lancet 358, 221–229 (2001).

38. Trepo, E., Romeo, S., Zucman-Rossi, J. & Nahon, P. PNPLA3 gene in liver diseases. J. Hepatol. 65, 399–412 (2016).

39. Xu, R., Tao, A., Zhang, S., Deng, Y. & Chen, G. Association between patatin-like phospholipase domain containing 3 gene (PNPLA3) polymorphisms and nonalcoholic fatty liver disease: a HuGE review and meta-analysis. Sci. Rep. 5, 9284 (2015).

40. Simon, T.G. et al. Risk of severe COVID-19 and mortality in patients with established chronic liver disease: a nationwide matched cohort study. BMC Gastroenterol. 21, 439 (2021).

41. Grimaudo, S. et al. PNPLA3 and TLL-1 polymorphisms as potential predictors of disease severity in patients with COVID-19. Front. Cell Dev. Biol. 9, 627914 (2021).

42. Innes, H. et al. The rs738409 G allele in PNPLA3 is associated with a reduced risk of COVID-19 mortality and hospitalization. Gastroenterology 160, 2599–2601 (2021).

43. Thomas, D.L. et al. Genetic variation in IL28B and spontaneous clearance of hepatitis C virus. Nature 461, 798–801 (2009).

44. Rauch, A. et al. Genetic variation in IL28B is associated with chronic hepatitis C and treatment failure: a genome-wide association study. Gastroenterology 138, 1338–1345 (2010).

45. Saponi-Cortes, J.M.R. et al. IFNL4 genetic variant can predispose to COVID-19. Sci. Rep. 11, 21185 (2021).

46. Agwa, S.H.A. et al. Association between interferon-lambda-3 rs12979860, TLL1 rs17047200 and DDR1 rs4618569 variant polymorphisms with the course and outcome of SARS-CoV-2 patients. Genes (Basel) 12, 830 (2021).

47. Balaouras, G., Eusebi, P. & Kostoulas, P. Systematic review and meta-analysis of the effect of ABO blood group on the risk of SARS-CoV-2 infection. PLoS One 17, e0271451 (2022).

48. Bshaena, A.M. et al. Association between ABO blood group system and COVID-19 severity. Am. J. Clin. Pathol., aqac106 (2022).

49. Kandasamy, K. et al. NetPath: a public resource of curated signal transduction pathways. Genome Biol. 11, R3 (2010).

50. Yamamoto, S. et al. INOH: ontology-based highly structured database of signal transduction pathways. Database (Oxford) 2011, bar052 (2011).

51. Schaefer, C.F. et al. PID: the Pathway Interaction Database. Nucleic Acids Res. 37, D674–679 (2009).

52. Zhou, G. et al. NetworkAnalyst 3.0: a visual analytics platform for comprehensive gene expression profiling and meta-analysis. Nucleic Acids Res. 47, W234–W241 (2019).

53. Manichaikul, A. et al. Robust relationship inference in genome-wide association studies. Bioinformatics 26, 2867–2873 (2010).

54. Purcell, S. et al. PLINK: a tool set for whole-genome association and population-based linkage analyses. Am. J. Hum. Genet. 81, 559–575 (2007).

55. Carroll, R.J., Bastarache, L. & Denny, J.C. R PheWAS: data analysis and plotting tools for phenome-wide association studies in the R environment. Bioinformatics 30, 2375–2376 (2014).

